# Understanding the patterns of repeated testing for COVID-19: Association with patient characteristics and outcomes

**DOI:** 10.1101/2020.07.26.20162453

**Authors:** Stephen Salerno, Zhangchen Zhao, Swaraaj Prabhu Sankar, Maxwell Salvatore, Tian Gu, Lars G. Fritsche, Seunggeun Lee, Lynda D. Lisabeth, Thomas S. Valley, Bhramar Mukherjee

## Abstract

**Importance:** The diagnostic tests for COVID-19 have a high false negative rate, but not everyone with an initial negative result is re-tested. Michigan Medicine, being one of the primary regional centers accepting COVID-19 cases, provided an ideal setting for studying COVID-19 repeated testing patterns during the first wave of the pandemic.

**Objective:** To identify the characteristics of patients who underwent repeated testing for COVID-19 and determine if repeated testing was associated with patient characteristics and with downstream outcomes among positive cases.

**Design:** This cross-sectional study described the pattern of testing for COVID-19 at Michigan Medicine. The main hypothesis under consideration is whether patient characteristics differed between those tested once and those who underwent multiple tests. We then restrict our attention to those that had at least one positive test and study repeated testing patterns in patients with severe COVID-19 related outcomes (testing positive, hospitalization and ICU care).

**Setting:** Demographic and clinical characteristics, test results, and health outcomes for 15,920 patients presenting to Michigan Medicine between March 10 and June 4, 2020 for a diagnostic test for COVID-19 were collected from their electronic medical records on June 24, 2020. Data on the number and types of tests administered to a given patient, as well as the sequences of patient-specific test results were derived from records of patient laboratory results.

**Participants:** Anyone tested between March 10 and June 4, 2020 at Michigan Medicine with a diagnostic test for COVID-19 in their Electronic Health Records were included in our analysis.

**Exposures:** Comparison of repeated testing across patient demographics, clinical characteristics, and patient outcomes

**Main Outcomes and Measures:** Whether patients underwent repeated diagnostic testing for SARS CoV-2 in Michigan Medicine

**Results:** Between March 10th and June 4th, 19,540 tests were ordered for 15,920 patients, with most patients only tested once (13596, 85.4%) and never testing positive (14753, 92.7%). There were 5 patients who got tested 10 or more times and there were substantial variations in test results within a patient. After fully adjusting for patient and neighborhood socioeconomic status (NSES) and demographic characteristics, patients with circulatory diseases (OR: 1.42; 95% CI: (1.18, 1.72)), any cancer (OR: 1.14; 95% CI: (1.01, 1.29)), Type 2 diabetes (OR: 1.22; 95% CI: (1.06, 1.39)), kidney diseases (OR: 1.95; 95% CI: (1.71, 2.23)), and liver diseases (OR: 1.30; 95% CI: (1.11, 1.50)) were found to have higher odds of undergoing repeated testing when compared to those without. Additionally, as compared to non-Hispanic whites, non-Hispanic blacks were found to have higher odds (OR: 1.21; 95% CI: (1.03, 1.43)) of receiving additional testing. Females were found to have lower odds (OR: 0.86; 95% CI: (0.76, 0.96)) of receiving additional testing than males. Neighborhood poverty level also affected whether to receive additional testing. For 1% increase in proportion of population with annual income below the federal poverty level, the odds ratio of receiving repeated testing is 1.01 (OR: 1.01; 95% CI: (1.00, 1.01)).

Focusing on only those 1167 patients with at least one positive result in their full testing history, patient age in years (OR: 1.01; 95% CI: (1.00, 1.03)), prior history of kidney diseases (OR: 2.15; 95% CI: (1.36, 3.41)) remained significantly different between patients who underwent repeated testing and those who did not. After adjusting for both patient demographic factors and NSES, hospitalization (OR: 7.44; 95% CI: (4.92, 11.41)) and ICU-level care (OR: 6.97; 95% CI: (4.48, 10.98)) were significantly associated with repeated testing. Of these 1167 patients, 306 got repeated testing and 1118 tests were done on these 306 patients, of which 810 (72.5%) were done during inpatient stays, substantiating that most repeated tests for test positive patients were done during hospitalization or ICU care. Additionally, using repeated testing data we estimate the “real world” false negative rate of the RT-PCR diagnostic test was 23.8% (95% CI: (19.5%, 28.5%)).

**Conclusions and Relevance:** This study sought to quantify the pattern of repeated testing for COVID-19 at Michigan Medicine. While most patients were tested once and received a negative result, a meaningful subset of patients (2324, 14.6% of the population who got tested) underwent multiple rounds of testing (5,944 tests were done in total on these 2324 patients, with an average of 2.6 tests per person), with 10 or more tests for five patients. Both hospitalizations and ICU care differed significantly between patients who underwent repeated testing versus those only tested once as expected. These results shed light on testing patterns and have important implications for understanding the variation of repeated testing results within and between patients.

**Key Points:** *Question:* Does having repeated diagnostic tests for the novel coronavirus (COVID-19) depend on patient characteristics and disease outcomes?

*Findings:* This cross-sectional study of testing patterns with 15,920 patients tested for SARS-CoV-2 virus at Michigan Medicine found significant differences in testing rates across patient age, body mass index, sex, race/ethnicity, neighborhood poverty level, prior history of circulatory diseases, any cancer, Type 2 diabetes, kidney, and liver diseases. Higher hospitalization rates and intensive care unit admissions were associated with repeated testing as expected.

*Meaning:* The results of this study describe diagnostic testing patterns for the novel COVID-19 virus at Michigan Medicine, and how they relate to patient characteristics and COVID-19 outcomes.

## Introduction

On March 10, 2020, the first two positive novel coronavirus (COVID-19) cases in the state of Michigan were identified by officials at the Department of Health and Human Services.^1^ A state of emergency was promptly declared on March 10, and a Gubernatorial stay-at-home order went into effect on March 23rd.^2^ In the ensuing months, a deluge of information on state-wide case counts had been reported.^3^ By necessity, such case reporting reflects a non-random sample of the infection status in tested individuals, leaving a large unknown regarding asymptomatic and untested infections. Thus, interpretation of the reported case-count data must be taken in the context of testing strategies and patterns to better understand the true extent of this disease. Moreover, the diagnostic tests had poor sensitivity (reported between 70-85%).^4^ As such, it is important to understand who got tested for COVID-19 and in particular, who got tested multiple times. The availability of diagnostic tests and guidelines also changed over time, with gradual relaxation of testing criteria and a large-scale expansion on May 26 prior to reopening the economy in the state of Michigan.^5^ Michigan Medicine, being one of the primary regional centers accepting COVID-19 cases from throughout the state, provided an ideal setting for studying COVID-19 testing patterns.

In this paper, we are particularly interested in characterizing who got tested more than once. Repeated testing needs to be considered in the clinical context of why tests were repeated. There are four major clinical reasons that may prompt a repeated test. First, there are individuals with high pretest probability of disease who have initial test results that are negative, prompting retest. Second, there are individuals who develop COVID symptoms and test negative initially and then may develop COVID symptoms again later and get tested again. Third, there are individuals who test positive and then get a repeat test to demonstrate that they are negative (since the CDC guidelines suggest two pathways for ending self-isolation: a) self-isolation for 10 days after testing or b) two negative tests separated by 24 hours).^6^ Fourth, there are individuals who test positive, are hospitalized, and then get a repeat test at the end of their disease course to confirm that they are now negative. Finally, frontline healthcare and essential workers get tested repeatedly. Thus, it is important to study the interval between two tests, the initial outcome of the test and whether the test was done inpatient or outpatient to understand repeated testing. This study helps us to compare and contrast what happened in practice with the recommended guidelines.^7^

Polymerase chain reaction (PCR) is a widely used COVID-19 diagnosis test. However, the sensitivity of PCR was 83.3% based on 36 patients finally diagnosed with COVID-19 at the Yichang Yiling Hospital,^8^ which indicates diagnosis tests may produce false negative results and diagnosis tests should be repeated to avoid misdiagnosis. Peto also pointed out the importance of repeated testing and advocates for its use as UK’s COVID-19 lockdown exit strategy.^9^ Several studies have already included repeated testing in their analysis.^10–12^ To the best of our knowledge, there are no studies exclusively focused on the repeated testing for COVID-19 as a an outcome with the goal of identifying patient characteristics that are associated with repeated testing.

Specifically, we examined associations between repeated testing and patient demographic, clinical characteristics, patient outcomes, differences between the types of tests administered, and patterns in return times and sequences of test results. The goals of this study were to (1) understand the pattern of repeated testing and the variation in test results for the same patient over time (2) identify the characteristics of patients who underwent repeated testing for COVID-19 and (3) determine if repeated testing was explained through COVID-19 outcomes (say hospitalization) among positive cases. Using results of repeated testing we also estimate empirical “real world” false negative rate of the test.

## Methods

### Study Sample

This cross-sectional study was approved by the committee for research ethics and compliance at Michigan Medicine and followed the Strengthening the Reporting of Observational Studies in Epidemiology (STROBE) reporting guidelines. Study protocols were reviewed and approved by the University of Michigan Medical School Institutional Review Board (IRB ID HUM00180294). All COVID-19 susceptible patients presenting to Michigan Medicine and tested between March 10 and June 4, 2020 were included in our analysis. Patient demographic and clinical characteristics, testing rates, test results, and health outcomes were collected from the electronic medical record (EMR) on June 24, 2020. Data on the number and types of tests administered to a given patient, as well as the sequences of patient-specific test results were derived from patient laboratory records. Seven different types of diagnostic tests were employed throughout the period of presentation, which differed both in terms of provider and type of specimen collected.

### Statistical Analyses

#### Frequency and Pattern of Testing

We describe the pattern of repeated testing for COVID-19 in our study population. We summarize the frequency of daily tests performed between March 10 and June 4, 2020, as well as the distribution of test results by the number of tests performed and the type of test administered. We examine the observed sequence of test results for those who were tested multiple times to characterize within subject variation in test results. We also calculate time to return of results, between two successive tests and for tests that were done inpatient or outpatient.

#### Association of Repeated Testing with Patient Characteristics

Bivariate tests are conducted to determine whether the distributions of several patient characteristics differed between those tested once and those tested multiple times. These characteristics included age (years), body mass index (BMI; kg/m^2^), sex (male, female, or other/unknown), race/ethnicity (non-Hispanic white, non-Hispanic black, or other/unknown), smoking status, and seven pre-existing comorbidities extracted from the electronic medical record: respiratory diseases, circulatory diseases, any cancers, Type 2 diabetes, kidney diseases, liver diseases, and autoimmune diseases. Each prevalent comorbidity was indicated as a binary predictor, 1: yes or 0: no. Neighborhood socioeconomic status (NSES) and demographic characteristics for each patient were derived from the National Neighborhood Data Archive (NaNDA) and included the proportion of the census tract population age 16+ in the civilian labor force who were unemployed, the proportion of the population with an annual income below the federal poverty level, and the proportion of adults with less than a high school diploma.^13^ Differences in these distributions are compared using chi-squared tests for discrete and Wilcoxon rank-sum tests for continuous variables.

Fully adjusted associations are tested by regressing an indicator of whether patients underwent repeated testing (1: tested more than once or 0: tested exactly once) as a binary outcome via a multivariable logistic regression model. We then restricted our study sample to include only those patients with a positive test result in their medical history. This subset of patients included those tested once and confirmed positive and those with at least one positive test result in a sequence of repeated tests. Updated unadjusted and adjusted comparisons were repeated for this sub-sample. We similarly restricted our analysis to hospitalized patients and patients needing ICU care to understand patient characteristics associated within patients with similar outcomes.

#### Association of Repeated Testing with COVID-19 Outcomes

We then explored whether patient prognosis was associated with repeated testing. Indicators for patient outcomes were considered sequentially in terms of severity. We first examined the relationship between the odds of repeated testing and a patient having ever tested positive. Of the patients with at least one positive test result, we then studied whether hospitalization (post COVID diagnosis), admission to the intensive care unit (ICU), or mortality were associated with repeated testing. To test these associations, we fit successive logistic regression models and obtained estimated odds ratios of patient prognosis. These odds ratios were reported as (a) unadjusted and (b) adjusted for age, sex, race/ethnicity, BMI, population density and smoking status. In a third level of adjustment (c), the NSES variables, neighborhood unemployment, poverty, and education levels, were incorporated into the model (b) to assess whether they confounded the relationships between repeated testing and each patient outcome. Finally, in fully adjusted models (d), we calculated these odds of repeated testing after controlling for (c) and a composite comorbidity score which was constructed by summing over the seven prevalent comorbidities indicators (0: no comorbidities to 7: all seven comorbidities).

Estimated Odds Ratios (OR) and Wald-type 95% confidence intervals were reported for all logistic regression models. A P-value <0.05 were noted for potential association.

### Sensitivity Analysis with Multiple Categories of Repeated Testing

In a sensitivity analysis, we then considered a further sub-classification for the patients who underwent repeated testing, in which we compared those tested once to those tested two to four times and those who underwent five or more tests. Stratified distributions for the patient and NSES characteristics by multiple testing categories were first studied for significant trends. We then fit a multinomial logistic regression model and a proportional odds model which directly contrasts both the odds of testing 2-4 times and the odds of testing 5+ times versus testing once. The results of this sensitivity analysis are presented in the supplementary material.

All analysis was carried out in R, version 4.0.0 (R Project for Statistical Computing).^14^

## Results

### Distributions of Testing Sequences

Figure 1 shows the daily patterns of COVID-19 testing, as well as aggregated summaries of repeated testing by test result and patient outcome. A total 15,920 patients presented to Michigan Medicine from March 10-June 4, 2020. During this period, 19,540 tests were ordered. As expected, weekly testing rates displayed a cyclical pattern, with more tests ordered during the week than on weekends. Additionally, there was an increase in testing going into the month of April, as there was increased community spread. It should be noted that testing policies changed from requiring an infectious disease consult to get tested to getting tested without a doctor’s note during this time period. In addition, there were seven types of diagnosis tests among all 19,540 tests. Most of them (18,947, 97%) came from COVID-19 PCR test (Figure S1). By and large, most patients were tested once (13596, 85.4%) and never tested positive (14753, 92.7%). Repeated testing occurred in a small, but meaningful, subset of the population (14.6%) with 10.2% of patients tested twice, 2.5% tested thrice, up to five patients undergoing ten or more repeated tests. Figure 2 displays the test results of these five patients with at least 10 tests, demonstrating the large variation of test results in the same patient. Four out of five patients ended with two negative consecutive tests, when self-isolation could be ended based on CDC guidelines. The other patient was still not discharged at time of the data pull. Among patients with at least one positive test result, 26.2% were tested more than once, as compared to 13.7% among those with all negative results. A greater proportion of hospitalized patients (47.6%), and the majority of patients needing an ICU care (61.7%), were tested more than once. Prior to March 17, uncertainty existed in the disease status of a small subset of patients (n = 78; 0.5%) who received presumptive test results. The majority of these 78 individuals received a single presumptive negative result (n = 54; 69.2%) or a single presumptive positive result (n = 10; 12.8%) and were not re-tested. Three patients (3.8%) received the same presumptive result in two consecutive tests and did not receive additional testing. The remaining 11 patients with an initial presumptive result (14.1%) were re-tested until they were confirmed negative (Table 1 & Table S1).

**Table 1:**
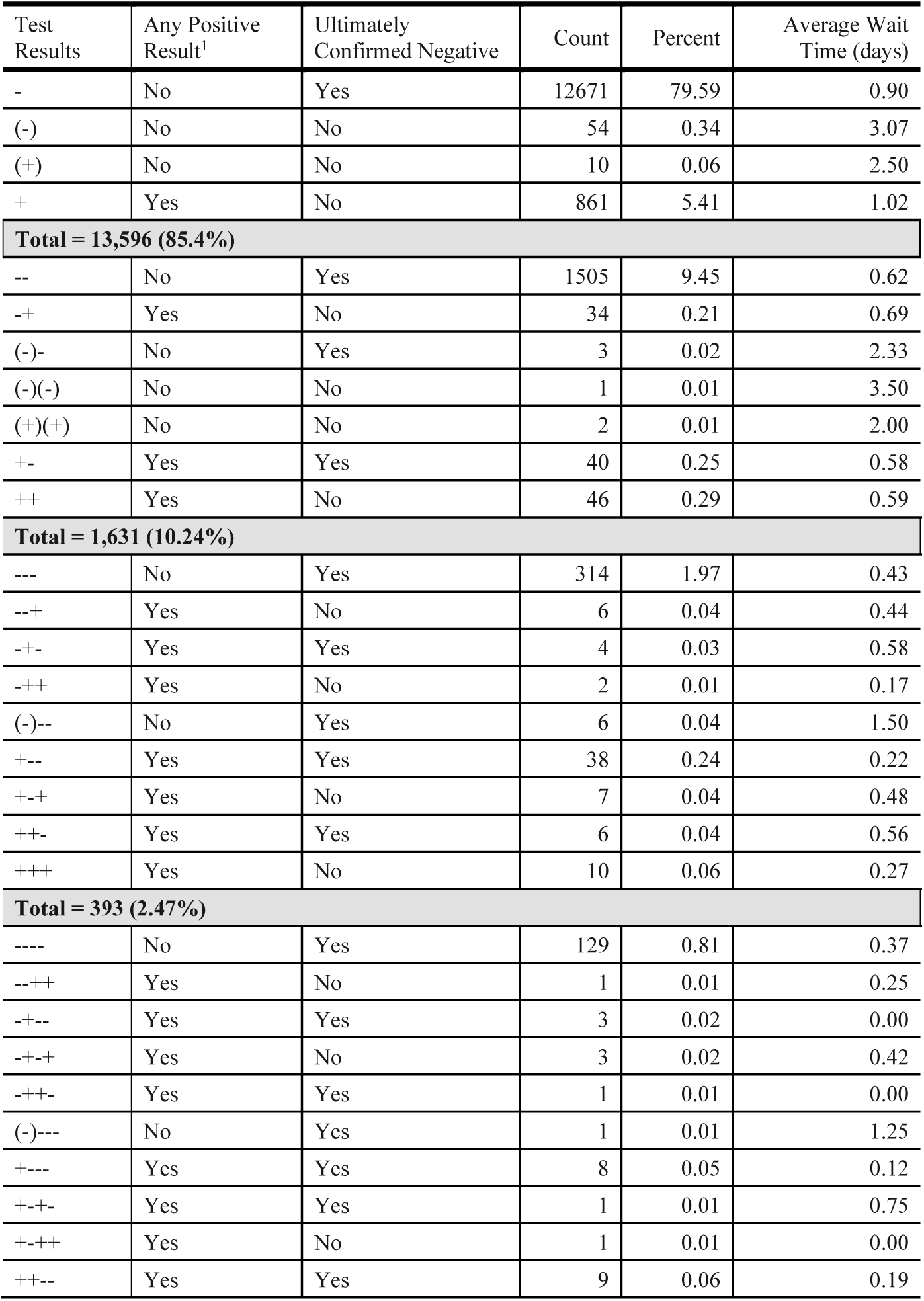
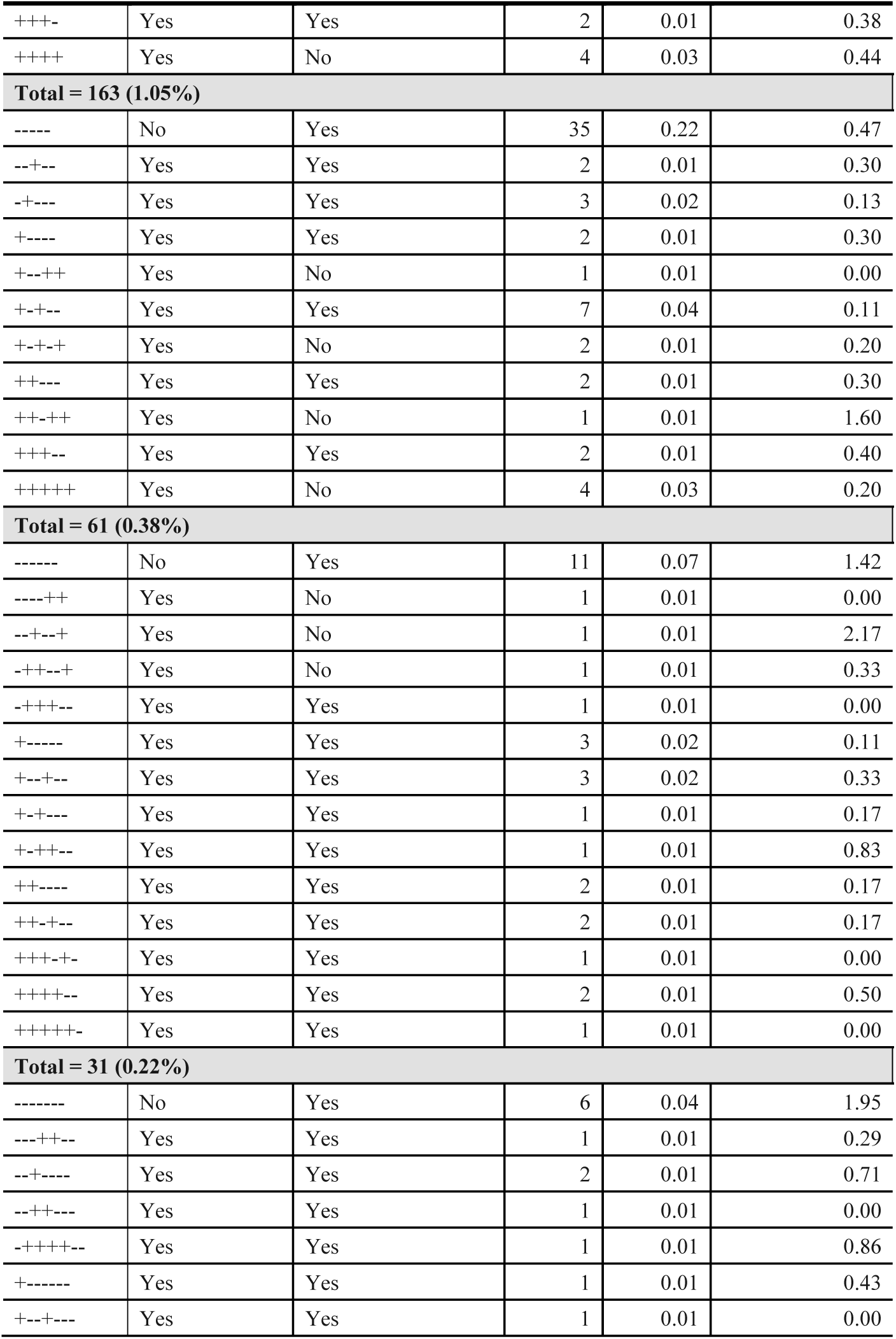
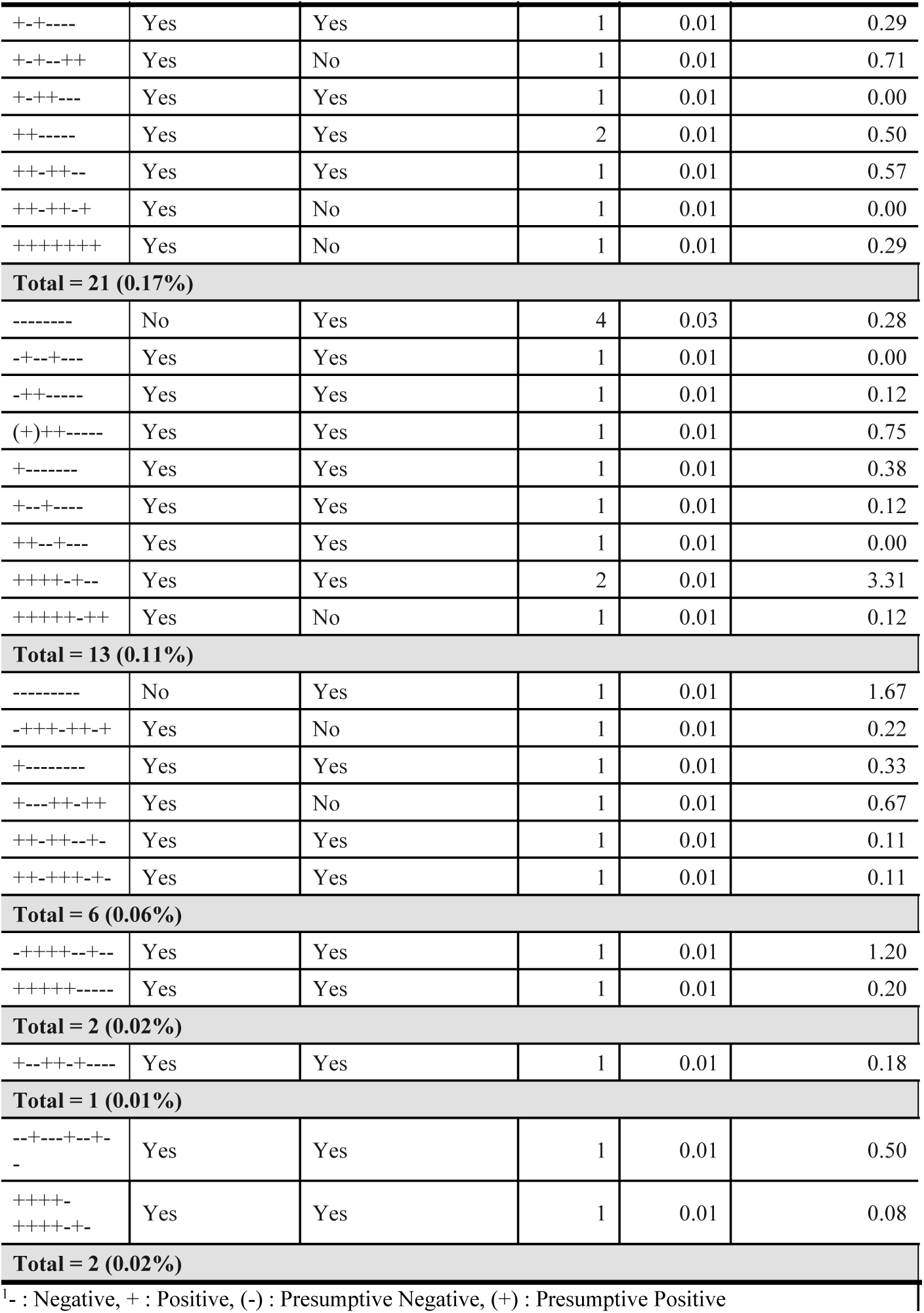
Sequences of testing results for the 19,540 tests ordered between March 10 and June 4, administered to the *n* = 15,920 patients presenting to Michigan Medicine before June 4, 2020.

**Figure 1:**
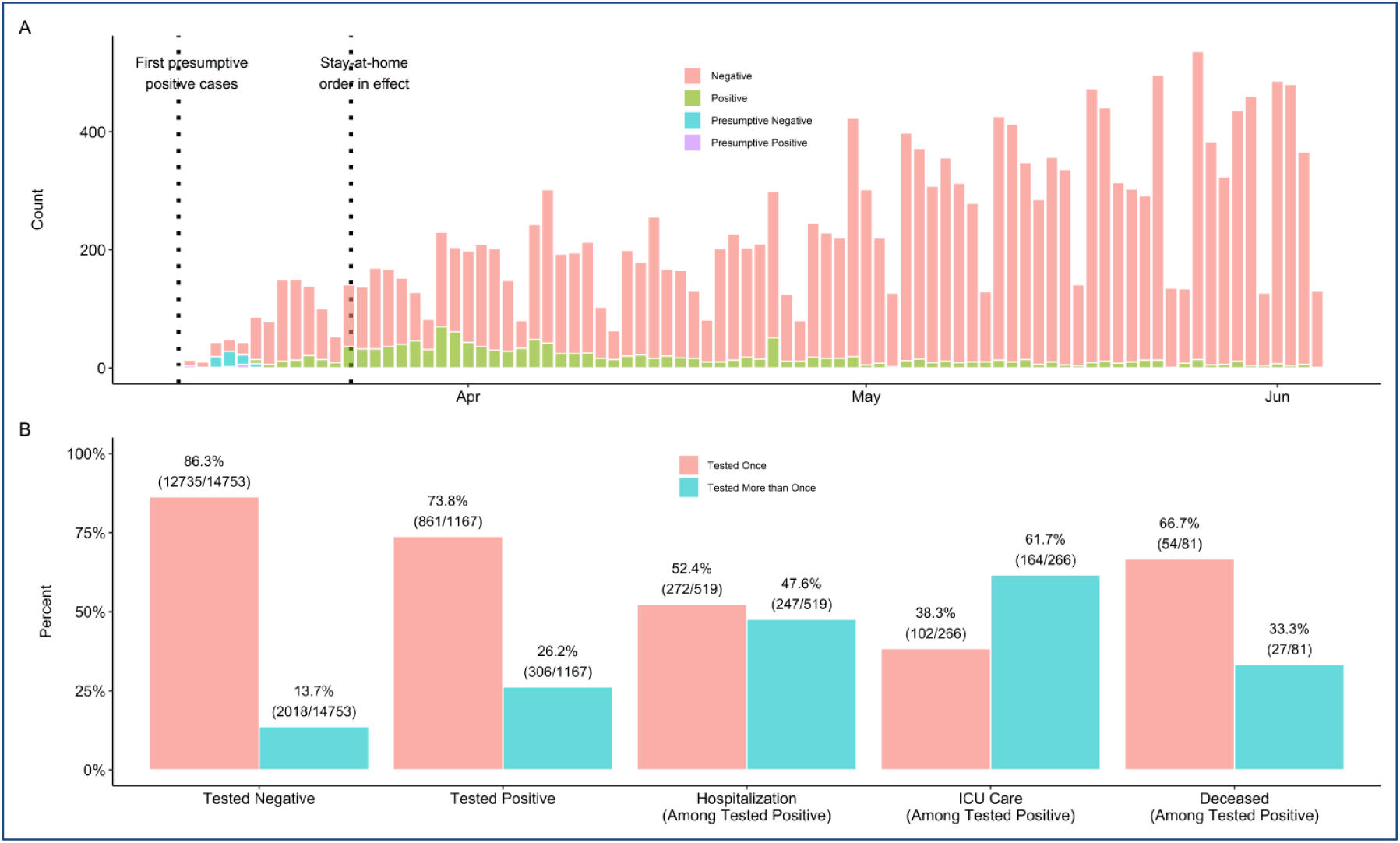
Patterns of COVID-19 testing at Michigan Medicine. (a) Number of tests by day, stratified by test result for the 15,920 patients presenting to Michigan Medicine before June 4, 2020. Between March 10th and June 4, 15,920 tests were ordered. (b) Proportion of patients tested once versus more than once among those testing negative, positive, hospitalized, admitted to the intensive care unit, and deceased.

**Figure 2:**
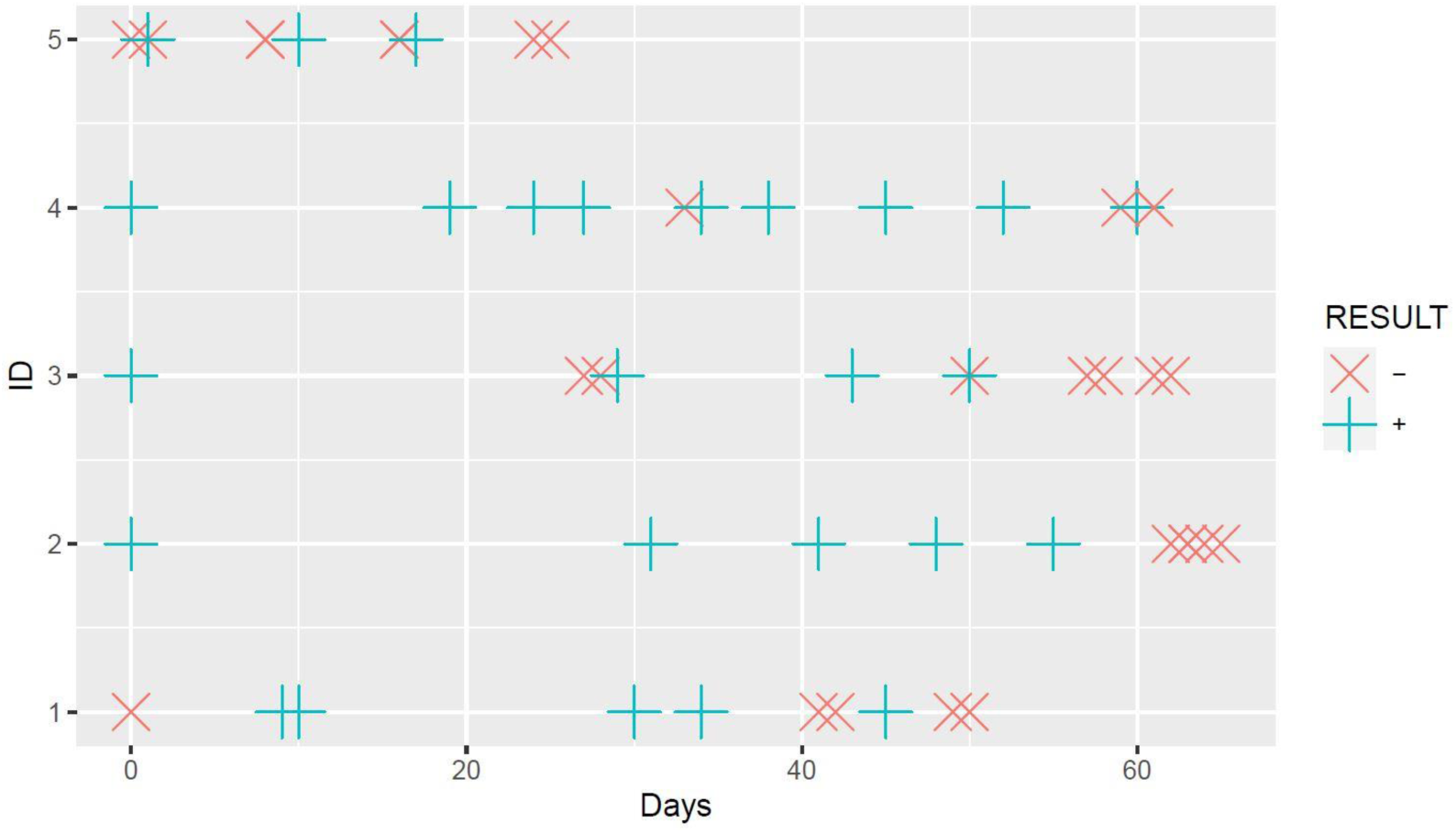
Test results of 5 patients who had at least 10 tests. The horizontal axis represents the test date (we marked the first test as day 0 for each patient) and the vertical axis represents patients’ ID.

For the 2342 patients who were tested more than once, 5,944 tests were ordered in total; on average, every patient had 2.6 (median: 25^th^ percentile: 2; 75^th^ percentile: 3; maximum: 12) tests. Across all sequences of results (Table 1) for those tested more than once, 13.2% received at least one positive result, but 94.3% ended up with an ultimately negative result. This compares to 6.4% testing positive among those patients tested once, thus showing repeated testing is associated with having an ultimate positive test. It is noteworthy how people with consistent sequence of negative results kept getting tested (314 had three successive negative tests, 129 had four successive negative tests 35 had five negative tests and 11 had a series of six negative tests). These could very likely be essential workers and health care professionals. The change in test results from an initial negative result to a positive result and the variation within a patient is surprising. For example, there were two patients with 12 test results with the result sequence --+---+--+-- and ++++-++++-+-, respectively (Figure 2). All 24 diagnosis used the same type of test, COVID-19 PCR test.

The average wait time for a test result was 13.8 hours (in hours: median: 3.67; min: 0.14.; 25^th^ percentile: 2.76; 75^th^ percentile: 17.6; max: 1514 hours) among patients tested repeatedly, which was significantly shorter than for those tested once (in hours: mean: 22.2; median: 18.2; min: 1.56; 25^th^ percentile: 3.22; 75^th^ percentile: 25.6; max: 1114; t: 12.22; P < 0.001). Among patients tested more than once, those with a positive test at any point experienced shorter average wait times for results (11.5 hours) as compared to those with all negative results (14.3 hours; t: 2.48; P =0.01). However, no significant differences were found between patients with an ultimately negative result (13.8 hours) and an ultimately positive result (13.5 hours; t: 0.20; P = 0.84) who were tested once.

Figure S2 shows distribution of days between two successive repeated tests. Among 15920 patients, 2324 patients had 2 or more tests; 693 patients had 3 or more tests; 300 patients had 4 or more tests and 137 had 5 or more tests. According to the current data, time gap between two successive tests appeared to be decreasing on average. The median time between 1^st^ and 2^nd^ test was 11 days, while the median time between 2^nd^ and 3^rd^ test decreased to 7 days. Both the median time between 3^rd^ and 4^th^ test and between 4^th^ and 5^th^ test was 6 days.

Lastly, there were 1979 tests conducted among 1167 patients with a positive test at any point. More than half of the tests (1053; 53.2%) were conducted on 519 patients during inpatient stays (Figure S3). Of these patients, 274 (52.8%) received one test, 65 (12.5%) were tested twice, 46 (8.9%) were tested thrice during inpatient stay, five patients (0.8%) who received ten or more inpatient tests. The inpatient tests had a shorter average wait times (8.64 hours) compared to outpatient tests (20.8 hours; t: 18.37; P < 0.001). As expected, patients undergoing repeated inpatient testing had significantly longer average stays (34.8 days) than those with one inpatient test (9.7 days; t: 9.63; P < 0.001). Among 519 patients with inpatient stays, 266 (51.3%) patients required ICU care (mean: 11.8 days; SD 10.3). Patients with ICU stays had significantly longer inpatient stays (28.8 days) than patients without ICU stays (7.88 days; t: 10.34; P<0.001). Specifically, 164 (61.7%) patients with ICU stay had repeated testing and had significantly longer inpatient stays (38.1 days) than 102 (38.3%) patients with ICU stay and only one test (15.5 days; t: 6.95; P<0.001).

### Associations between Repeated Testing and Patient Characteristics

Age, BMI, sex, race/ethnicity, smoking status, neighborhood unemployment, neighborhood poverty levels, neighborhood education and all seven prevalent comorbidities were found to differ significantly between patients tested once and those tested more than once (Table 2). After adjusting for all other demographic and clinical characteristics presented in Table 2, significant differences persisted with respect to patient age, BMI, sex, race/ethnicity, neighborhood poverty levels, prevalence of circulatory diseases, cancer and Type 2 diabetes, and indications of kidney and liver diseases. Specifically, patients with these prevalent comorbidities: circulatory diseases (OR: 1.42; 95% CI: (1.18, 1.72)), any cancer (OR: 1.14; 95% CI: (1.01, 1.29)), Type 2 diabetes (OR: 1.22; 95% CI: (1.06, 1.39)), kidney diseases (OR: 1.95; 95% CI: (1.71, 2.23)), and liver diseases (OR: 1.30; 95% CI: (1.11, 1.50)) were found to have higher odds of undergoing repeated testing when compared to those without. As compared to non-Hispanic whites, non-Hispanic blacks were found to have higher odds (OR: 1.21; 95% CI: (1.03, 1.43)) of receiving additional testing. In addition, females were found to have lower odds (OR: 0.86; 95% CI: (0.76, 0.96)) of receiving additional testing than males. Neighborhood poverty level also affected whether to receive additional testing. For 1% increase in proportion of population with annual income below the federal poverty level, the odds ratio of receiving repeated testing is 1.01 (OR: 1.01; 95% CI: (1.00, 1.01)). Lastly, age in years (OR: 1.01; 95% CI: (1.00, 1.01) and BMI (OR: 0.99; 95% CI: (0.98, 0.99)) had weak, but significant effects. Focusing on only those patients with at least one positive result in their full testing history, adjusting for all other demographic and clinical characteristics, significant differences were found with respect to patient age (OR: 1.01; 95% CI: (1.00, 1.03)) and indications of kidney diseases (OR: 2.15; 95% CI: (1.36, 3.41)). Only indications of kidney diseases (OR: 1.93; 95% CI: (1.05, 3.58)) remained significant if only including 519 patients with at least one positive result and hospital stays (Table S2). Additionally, no significant difference was found among 266 patients with at least one positive result and ICU stays.

**Table 2:** Characteristics of the *n* = 15,920 patients in our study sample and *n* = 1,167 patients with a positive COVID-19 test in their medical history, stratified by the number of tests underwent by the patient. Statistics presented are median (inter-quartile range) for continuous variables and n (%) for categorical variables. Unadjusted p-values are reported for either Wilcoxon rank-sum (continuous) or chi-square tests of independence (categorical) comparing the distributions of each of these characteristics between testing groups. Odds ratios and 95% confidence intervals are reported for each characteristic, fully adjusting for all other demographic and clinical characteristics in a logistic regression model.

| Characteristic | All Tested Patients: Unadjusted Comparisons |  |  |  | All Tested Patients: Logistic Regression |  |  | Patients with a Positive Test Result: Unadjusted Comparisons |  |  |  | Patients with a Positive Test Result: Logistic Regression |  |  |
| --- | --- | --- | --- | --- | --- | --- | --- | --- | --- | --- | --- | --- | --- | --- |
|  | Overall, N = 15920 | Tested Once, N = 13596<sup>1</sup> | Tested More Than Once, N = 2324<sup>1</sup> | p-value<sup>2</sup> | OR<sup>3</sup> | 95% CI<sup>3</sup> | p-value | Overall, N = 1167 | Tested Once, N = 861<sup>1</sup> | Tested More Than Once, N = 306<sup>1</sup> | p-value<sup>2</sup> | OR<sup>3</sup> | 95% CI<sup>3</sup> | p-value |
| Age, years | 49 (30 - 65) | 47 (29 - 64) | 57 (38 - 70) | **<0.001** | 1.01 | 1.00, 1.01 | **<0.001** | 53 (38 - 66) | 51 (37 - 64) | 59 (44 - 71) | **<0.001** | 1.01 | 1.00, 1.03 | **0.016** |
| Body Mass Index | 28 (24 - 33) | 28 (24 - 33) | 28 (24 - 33) | **0.015** | 0.99 | 0.98, 0.99 | **0.001** | 30 (26 - 36) | 30 (26 - 36) | 30 (25 - 36) | 0.71 | 1 | 0.97, 1.02 | 0.74 |
| Sex |  |  |  | **<0.001** |  |  |  |  |  |  | **0.016** |  |  |  |
| Male | 6767 (43) | 5639 (41) | 1128 (49) |  |  |  |  | 543 (47) | 382 (44) | 161 (53) |  |  |  |  |
| Female | 9153 (57) | 7957 (59) | 1196 (51) |  | 0.86 | 0.76, 0.96 | **0.008** | 624 (53) | 479 (56) | 145 (47) |  | 0.75 | 0.52, 1.10 | 0.14 |
| Race/Ethnicity |  |  |  | **<0.001** |  |  |  |  |  |  | **0.033** |  |  |  |
| Non-Hispanic White | 11402 (72) | 9804 (72) | 1598 (69) |  |  |  |  | 527 (45) | 387 (45) | 140 (46) |  |  |  |  |
| Non-Hispanic Black | 2136 (13) | 1735 (13) | 401 (17) |  | 1.21 | 1.03, 1.43 | **0.021** | 409 (35) | 289 (34) | 120 (39) |  | 0.9 | 0.58, 1.39 | 0.63 |
| Other/Unknown | 2382 (15) | 2057 (15) | 325 (14) |  | 1.05 | 0.88, 1.26 | 0.57 | 231 (20) | 185 (21) | 46 (15) |  | 0.7 | 0.36, 1.29 | 0.27 |
| Smoking Status |  |  |  | **<0.001** |  |  |  |  |  |  | 0.14 |  |  |  |
| Current/Former | 5856 (37) | 4863 (36) | 993 (43) |  |  |  |  | 339 (29) | 237 (28) | 102 (33) |  |  |  |  |
| Never | 8904 (56) | 7733 (57) | 1171 (50) |  | 0.96 | 0.86, 1.08 | 0.52 | 626 (54) | 474 (55) | 152 (50) |  | 0.95 | 0.65, 1.40 | 0.8 |
| Unknown | 1160 (7.3) | 1000 (7.4) | 160 (6.9) |  | 0.95 | 0.50, 1.67 | 0.87 | 202 (17) | 150 (17) | 52 (17) |  | 1.15 | 0.33, 3.44 | 0.82 |
| Neighborhood Unemployment<sup>4</sup> | 6 (4 - 8) | 6 (4 - 8) | 6 (4 - 9) | **0.016** | 1.00 | 0.98, 1.03 | 0.65 | 7 (5 - 9) | 7 (5 - 10) | 7 (4 - 9) | 0.7 | 1.01 | 0.95, 1.07 | 0.85 |
| Neighborhood Poverty<sup>4</sup> | 9 (5 - 17) | 8 (4 - 17) | 10 (6 - 19) | **<0.001** | 1.01 | 1.00, 1.01 | **0.039** | 10 (6 - 20) | 10 (5 - 19) | 12 (6 - 22) | **0.038** | 1.02 | 0.99, 1.04 | 0.13 |
| Neighborhood Education<sup>4</sup> | 7 (4 - 11) | 6 (4 - 11) | 7 (4 - 12) | **0.001** | 1.00 | 0.99, 1.01 | 0.97 | 8 (4 - 13) | 8 (4 - 13) | 8 (4 - 13) | 0.75 | 1.00 | 0.96, 1.03 | 0.86 |
| Population Density (per 100 people) | 20.6 (5.2 - 36.4) | 20.1 (5.1 - 36.1) | 21.7 (6.0 - 38.6) | **0.04** | 1 | 1.00, 1.00 | 0.9 | 27.4 (12.4 - 47.0) | 27.4 (12.2 - 45.5) | 29.1 (14.0 - 47.6) | 0.35 | 1 | 1.00, 1.00 | 0.68 |
| Respiratory Diseases |  |  |  | **<0.001** |  |  |  |  |  |  | 0.2 |  |  |  |
| No | 3633 (26) | 3256 (27) | 377 (19) |  |  |  |  | 208 (25) | 163 (26) | 45 (21) |  |  |  |  |
| Yes | 10541 (74) | 8881 (73) | 1660 (81) |  | 1.07 | 0.91, 1.25 | 0.41 | 637 (75) | 469 (74) | 168 (79) |  | 0.76 | 0.47, 1.25 | 0.28 |
| Circulatory Diseases |  |  |  | **<0.001** |  |  |  |  |  |  | **<0.001** |  |  |  |
| No | 3787 (27) | 3487 (29) | 300 (15) |  |  |  |  | 220 (26) | 185 (29) | 35 (16) |  |  |  |  |
| Yes | 10387 (73) | 8650 (71) | 1737 (85) |  | 1.42 | 1.18, 1.72 | **<0.001** | 625 (74) | 447 (71) | 178 (84) |  | 1.59 | 0.91, 2.88 | 0.11 |
| Any Cancer |  |  |  | **<0.001** |  |  |  |  |  |  | **0.01** |  |  |  |
| No | 9523 (67) | 8358 (69) | 1165 (57) |  |  |  |  | 615 (73) | 475 (75) | 140 (66) |  |  |  |  |
| Yes | 4651 (33) | 3779 (31) | 872 (43) |  | 1.14 | 1.01, 1.29 | **0.03** | 230 (27) | 157 (25) | 73 (34) |  | 1.17 | 0.78, 1.76 | 0.44 |
| Type 2 Diabetes |  |  |  | **<0.001** |  |  |  |  |  |  | **<0.001** |  |  |  |
| No | 11431 (81) | 9994 (82) | 1437 (71) |  |  |  |  | 629 (74) | 493 (78) | 136 (64) |  |  |  |  |
| Yes | 2743 (19) | 2143 (18) | 600 (29) |  | 1.22 | 1.06, 1.39 | **0.004** | 216 (26) | 139 (22) | 77 (36) |  | 1.19 | 0.77, 1.84 | 0.43 |
| Kidney Diseases |  |  |  | **<0.001** |  |  |  |  |  |  | **<0.001** |  |  |  |
| No | 11370 (80) | 10079 (83) | 1291 (63) |  |  |  |  | 663 (78) | 532 (84) | 131 (62) |  |  |  |  |
| Yes | 2804 (20) | 2058 (17) | 746 (37) |  | 1.95 | 1.71, 2.23 | **<0.001** | 182 (22) | 100 (16) | 82 (38) |  | 2.15 | 1.36, 3.41 | **0.001** |
| Liver Diseases |  |  |  | **<0.001** |  |  |  |  |  |  | 0.27 |  |  |  |
| No | 12612 (89) | 10923 (90) | 1689 (83) |  |  |  |  | 764 (90) | 576 (91) | 188 (88) |  |  |  |  |
| Yes | 1562 (11) | 1214 (10) | 348 (17) |  | 1.3 | 1.11, 1.50 | **<0.001** | 81 (9.6) | 56 (8.9) | 25 (12) |  | 1.04 | 0.58, 1.82 | 0.89 |
| Autoimmune Diseases |  |  |  | **<0.001** |  |  |  |  |  |  | 0.05 |  |  |  |
| No | 11580 (82) | 10008 (82) | 1572 (77) |  |  |  |  | 665 (79) | 508 (80) | 157 (74) |  |  |  |  |
| Yes | 2594 (18) | 2129 (18) | 465 (23) |  | 1.03 | 0.90, 1.18 | 0.64 | 180 (21) | 124 (20) | 56 (26) |  | 1.24 | 0.81, 1.89 | 0.32 |

<sup>1</sup>Statistics presented: median (IQR); n (%)
<sup>2</sup>Statistical tests performed: Wilcoxon rank-sum test; chi-square test of independence
<sup>3</sup>OR = Odds Ratio, CI = Confidence Interval
<sup>4</sup>The unit of neighborhood unemployment is 1% proportion of population age 16+ in the civilian labor force who are unemployed; the unit of neighborhood poverty is 1% proportion of population with annual income below the federal poverty level; and the unit of neighborhood education is 1% proportion of adults with less than high school diploma in 2010.

Our sensitivity analysis revealed that patient demographics and NSES further differed across repeated testing groups (tested 2-4 times and tested 5+ times versus tested once). Results from our fully adjusted multinomial model (Table S3) revealed that, in addition to patient age, BMI, sex, race/ethnicity, neighborhood poverty levels, and prevalence of circulatory diseases, any cancer, Type 2 diabetes, kidney and liver diseases, smoking status, neighborhood unemployment, neighborhood education levels, and indications of respiratory diseases, and autoimmune diseases were also significantly associated with repeated testing. Respiratory diseases (tested 2-4 times versus once: (OR: 1.04; 95% CI: (1.00, 1.08)); tested 5+ times versus once (OR: 1.76; 95% CI: (1.76, 1.76))), and autoimmune diseases (tested 5+ times versus once (OR: 1.42; 95% CI: (1.42, 1.43))) were found to have higher odds of undergoing repeated testing when compared to those without. In addition, higher neighborhood unemployment were associated with higher odds of repeated testing (tested 2-4 times versus one: (OR: 1.01; 95% CI: (1.01, 1.01)); tested 5+ times versus once: (OR: 1.01; 95% CI: (1.01, 1.01)) per 1% increase in proportion of population. The results remained consistent when considering a proportional odds model instead, and when restricting the patient population to only those patients with a positive test in their medical history.

### Associations between Repeated Testing and Patient Outcomes

Among 2324 patients underwent repeated testing, 812 out of 5944 tests (13.7%) were done during in patient stays. Thus, most repeated tests were done outpatient. If we focus on the patients with repeated tests and at least one positive result (N=306), then 49 out of 1118 tests (4.38%) were done before hospitalization and 810 out of 1118 tests (72.5%) were done during inpatient stays substantiating most repeated tests were done during hospitalization or ICU care for patients that tested positive. This finding confirms one of the reasons for repeated tests: patients at hospital need test more to confirm they are negative and disease free.

Table 3 displays the odds ratios for repeated testing for three successive patient outcomes: positive test, hospitalization, ICU requirement. Under unadjusted framework, testing positive, being hospitalized, and requiring ICU-level care are significantly associated with repeated testing. After three sets of adjustments for both patient characteristics and NSES, the odds of repeated testing remained significantly higher for patients who being tested positive (OR: 2.00; 95% CI: (1.66, 2.39)); being hospitalized (OR: 7.44; 95% CI: (4.92, 11.41)) and requiring ICU-level care (OR: 6.97; 95% CI: (4.48, 10.98)) than patients without these outcomes.

**Table 3:** Associations between repeated testing and patient outcomes. Statistics presented are odds ratios and 95% confidence intervals for the associations of each patient outcome with repeated testing from logistic regression models that are (a) unadjusted, (b) adjusted for age, sex at birth, race, ethnicity, and neighborhood population density, (c) adjusted for (b) and the proportion of adults with less than a high school education, in the labor force but unemployed, with income below the federal poverty level, and (d) adjusted for (c) and the composite comorbidity score. All OR correspond to the odds of getting repeated tested versus tested once.

| Adjustment Model | Test Positive<br>(Among All Tested) | Hospitalization<br>(Among Tested<br>Positive) | ICU Care<br>(Among Tested<br>Positive) | Deceased<br>(Among Tested<br>Positive) |
| --- | --- | --- | --- | --- |
| (a) unadjusted | **2.24 (1.95, 2.57)** | **8.88 (6.51, 12.27)** | **8.48 (6.27, 11.54)** | 1.39 (0.84, 2.24) |
| (b) adjusted for age, sex at birth, race, ethnicity, and neighborhood population density | **1.82 (1.52, 2.16)** | **7.88 (5.29, 11.93)** | **6.74 (4.43, 10.34)** | 0.87 (0.43, 1.70) |
| (c) adjusted for (b) and the proportion of adults with less than a high school education, in the labor force but unemployed, with income below the federal poverty level | **1.83 (1.54, 2.18)** | **8.12 (5.42, 12.36)** | **7.06 (4.61, 10.94)** | 0.84 (0.41, 1.65) |
| (d) adjusted for (c) and the composite comorbidity score | **2.00 (1.66, 2.39)** | **7.44 (4.92, 11.41)** | **6.97 (4.48, 10.98)** | 0.74 (0.35, 1.50) |

Our sensitivity analysis modeled the associations of each patient outcome with repeated testing group as a factor (tested 2-4 times and tested 5+ times versus tested once) from multinomial logistic regression models (Table S4). After adjustments, the odds of being tested 2-4 times for patients testing positive (OR: 1.55; 95% CI: (1.54, 1.55)); being hospitalized (OR: 5.62; 95% CI: (4.68, 6.76)) and requiring ICU-level care (OR: 4.61; 95% CI: (4.08, 5.21)) were significantly higher than patients without outcomes. Additionally, the odds of 5+ tests for patients being positive; being hospitalized and requiring ICU-level care were even higher than the odds of 2-4 tests as expected.

## Discussion

Understanding testing rates for COVID-19 is necessary to garner insight about the scope and severity of this disease, beyond what is ascertainable from case counts alone. We have shown that the vast majority of patients at Michigan Medicine were tested once, and that this test resulted in a negative result. A meaningful subset of patients (N=2324) underwent multiple rounds of testing. Among these patients, most patients (N=2070; 89.1%) received consistent diagnosis; 166 (7.1%) patients received one flip (from + to -;or from – to +) during repeated tests; 81 (3.5%) patients received more than one flip, demonstrating the high variation in the testing results, which is consistent with high false negative rate in the current testing technology. We can also make a rough estimate of the false negative rate from repeated testing data. There were 85 patients with at least two positive tests and at least one negative test between two positive tests (total tests=370). Due to low false positive rate of the RT-PCR test (0.8-4%),^15^ we assume that all test results should remain positive between two positive tests. Under this assumption, there were 88 negative tests that should be positive and then the estimation of false negative rate was (88/370) or 23.8% (95% CI: (19.5%, 28.5%)).

At Michigan Medicine, 319 out of 15,920 patients tested by RT-PCR have already conducted serology test for presence of IgG antibody (Table S5). **Among patients with all negative diagnostic tests, patients with one diagnosis test had significant chance to test positive in serology tests compared to patients with repeated diagnosis tests, which implied both the high false negative rate in diagnosis tests and the necessity of repeated testing**. Specifically, 37 (18.7%) patients with only one negative diagnosis test had positive serology test result, which was close to the false negative rate 23.8% we estimated before. If repeated tests are assumed to be independent and have 18.7% false negative rate, then the probability to get two false negative results is as low as 3.50%. However, 6 (14.3%) patients with repeated negative diagnosis tests had positive serology test result. There are three reasons that might explain the difference. First, patients might be infected with COVID-19 between two diagnosis tests (i.e. only one false negative); second, patients might be infected between diagnosis tests and serology tests (i.e. no false negative in diagnosis tests); third, our assumption of independence may be violated, and some patients may have higher change to receive false negative results. Overall, from the serology data, it appears that repeated diagnosis testing can effectively avoid the severe consequences from high false negative rate in diagnosis tests and help reduce the spread of COVID-19.

We demonstrated that testing rates differed significantly with respect to age, BMI, sex, race/ethnicity, neighborhood poverty and indications of circulatory diseases, any cancer, Type 2 diabetes, kidney, and liver diseases. Among those patients with a positive test result in their medical record, differences in testing rates persisted with respect to age, and an indication of prevalent kidney diseases. The variation in COVID-19 results provide an idea about the accuracy of the tests and the desire to get repeatedly tested.

Considering patient prognosis being associated with repeated testing or leading to repeated testing, we found that the odds of repeated testing were significantly higher for patients that were hospitalized or required ICU stays, adjusting for patient characteristics and NSES. It is most likely that patients who were admitted to the hospital or who had an ICU requirement were tested before release if they were returning to homes/assisted living where they could not practice self-isolation. The weak association of mortality with repeated testing may also indicate that patients with extremely severe symptoms were less likely to get repeated tests. Inpatient stays lasted more than three weeks, on average, among those patients repeatedly tested. Therefore, unlike a normal flu, it took much longer than expected to recover from COVID-19 and get a stable negative test result.^16–18^

Countries like South Korea have shown that random testing and contact tracing are crucial in understanding the true prevalence of this disease and managing its spread.^19^ The United States was initially not prepared to reinforce such testing strategies at the onset of the domestic outbreak.^20^ Initial testing guidelines called for the rationing of resources, whereby only the most critically ill patients or those at highest risk of presenting with severe symptoms were tested. This potentially delayed the “flattening of the curve” strategy, as disease transmission continued to occur between asymptomatic individuals. At Michigan Medicine, more and more asymptomatic individuals got tests in May and June because more tests were available. As of July 23, 51,680,022 COVID-19 tests have been reported in the US (15.74 tests per 100 people) and 1,634,670 COVID-19 tests have been reported in Michigan (16.37 tests per 100 people).^21,22^ As more people got tested and lockdown policy kept extended by Michigan governor, both daily new cases and positive rates kept decreasing.

## Limitations

A potential limitation of our study is its generalizability to other regional testing centers, both in established “hot spots” and rural areas, due to the patient mix at Michigan Medicine. There are inherent limitations to using electronic health records for research purposes due to the incomplete information. For example, tests done at drive-thru testing stations or pharmacies are not captured. The definition of co-morbidities and patient characteristics using ICD code can be highly imperfect.

## Conclusions

This study sought to quantify patterns of repeated testing for COVID-19 and its associated factors at Michigan Medicine. These results shed light on testing patterns and have important implications in understanding what is happening in real world with COVID testing in an academic medical center. It also gives a real world estimate of the false negative rate of the test.

## Data Availability

Data cannot be shared publicly due to patient confidentiality. The data underlying the results presented in the study are available from University of Michigan Medical School Central Biorepository for researchers who meet the criteria for access to confidential data.

